# Dual-domain joint learning reconstruction method (JLRM) combined with physical process for spectral computed tomography

**DOI:** 10.1101/2024.01.22.24301600

**Authors:** Genwei Ma, Xing Zhao

## Abstract

Spectral computed tomography (SCT) is an powerful imaging modality with broad applications and advantages such as contrast enhancement, artifact reduction, and material differentiation. The positive process or data collected process of SCT is a nonlinear physical process existing scatter and noise, which make it is an extremely ill-posed inverse problem in mathematics. In this paper, we propose a dual-domain iterative network combining a joint learning reconstruction method (JLRM) with a physical process. Specifically, a physical module network is constructed according to the SCT physical process to accurately describe this forward process, which makes the nonlinear use of the traditional mathematical iterative algorithm effective and stable. Additionally, we build a residualto-residual strategy with an attention mechanism to overcome the slow speed of the traditional mathematical iterative algorithm. We have verified the feasibility of the method through our winning submission to the AAPM DL-spectral CT challenge, and demonstrated that high-accuracy also basis material decomposition results can be achieved with noisy data.

## 1 Introduction

Spectral computed tomography (SCT) is popular and promising in clinical and industrial nondestructive testing. Traditional CT aims to accurately recover a detailed internal image of an object by scanning it from different angles. However, it does not consider the polychromaticity of x-rays. SCT, as shown in Fig. 1, considers the beam spectrum is broad and the polychromatic nature of the x-ray beam makes quantitative imaging. Considering the x-ray spectral information, this leads to more complex reconstruction problems. Hence, in SCT, an object is scanned with different x-ray spectra or spectral separations from the broad spectrum to supplement the information. Spectral x-ray data collection methods include kVp-switching [1], dual-source scanning [2], dual-layer detection, simplistic two-pass scanning, and photon counting detectors [3, 4]. Generally, the collected data of SCT, also known as polychromatic projection data, are utilized to perform energy- and material-selective reconstructions [5– 8]. In other words, SCT not only can distinguish materials better but allows quantification of the mass density of two or more materials in a tissue or a mixture with known elemental composition [9].

**Fig. 1.**
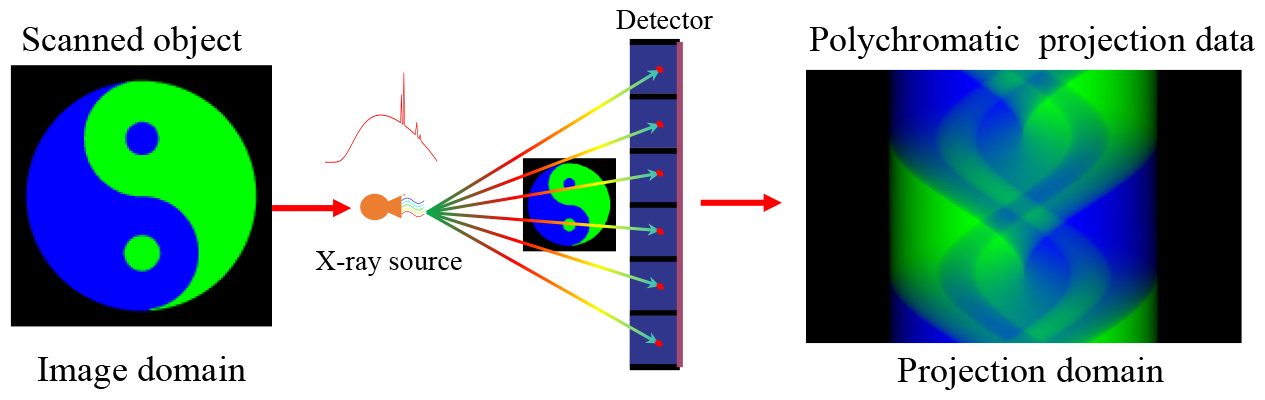
Spectral computed tomography imaging process.

In mathematics, the SCT reconstruction problem can be seen as solving for *µ*(*x, E*) from *K* sets of polychromatic projection data under different spectra or spectral separation,

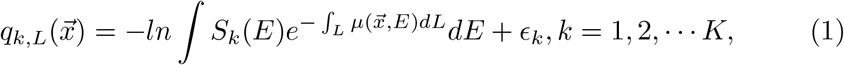

where 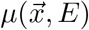 is the linear attenuation coefficient of the scanned object at point 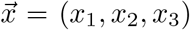 with energy *E* and *S*_*k*_(*E*), *k* = 1, 2.is normalized effective spectrum, ∫ *S*_*k*_(*E*)*dE* = 1 and *ϵ*_*k*_ represent scatter or noise of the *k*th projection. In SCT, the linear attenuation coefficient is generally decomposed as a linear combination of some predefined basis functions,

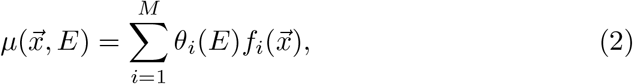

where *θ*_*i*_(*E*) are energy-dependent functions, and 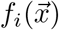 are functions of spatial position. There are two physical explanations for this equation. The first one is the decomposition method based on the physical effect, where *M* = 2 in this case and the functions based on the photoelectric effect and Compton scattering.The second explanation is basis material decomposition. *θ*_*i*_(*E*), represent the mass attenuation coefficients of the selected basis materials. The reconstructed 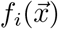, represent the densities of the basis materials of the scanned object.

Substituting (2) into (1), we can obtain the discrete form of SCT as follow:

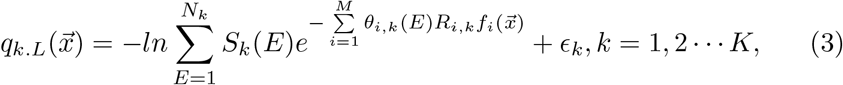

where *R*_*i,k*_, *k* = 1, 2, *K*, represents the *k*th Radon transform.

Compared with traditional CT, the reconstruction problem of SCT is nonlinear and ill-posed. Moreover, real projection data may be inconsistent. Algorithms to solve this problem can be classified in two ways. According to the domain where the data are processed, the methods can be divided into image domain methods [8–14] and projection domain methods [15, 16]. They can also be categorized into direct mapping methods [5, 10–15, 17–19], iterative algorithms [16, 20–31], and deep-learning methods [32–40]. Several researchers combine these classifications.

Image domain methods, also known as post-processing methods, directly process images that have been reconstructed from collected raw data. The reconstructed image is generally solved using a linear algorithm, such as the algebra reconstruction technique (ART) or filter back-projection (FBP). This is actually a first-order approximation of the original nonlinear data, although image domain methods use different basis functions [10, 11, 15, 18] to approximate it. Hence, the reconstructed images still suffer from artifacts such as beam-hardening. Projection domain methods process either raw or log-transformed data before image reconstruction. Theoretically, projection domain methods can obtain better images than image domain methods, because they can compensate for nonlinear effects on material decomposition and suppress artifacts in the decomposition process. However, polychromatic projections require geometric consistency, and it is notoriously difficult to practical SCT system. Direct mapping constructs a basis function and establishes a mapping between the SCT data and basis material. While direct mapping methods are quick and simple and do not consider the spectrum information and attenuation coefficient of materials, they are highly dependent on the calibration phantom, which limits the accuracy of the reconstructed images. Iteration reconstruction algorithms are the most effective way to solve ill-posed nonlinear inverse problems. Unfortunately, they are time-consuming when the geometric parameters of different spectral scans are inconsistent. Many researchers combine the statistical assumptions of raw data and prior information on images, but this involves hyperparameters that can only be adjusted empirically and do not offer consistent image quality improvement [41]. sourcedual-detector CT systems [42, 43]. In recent years, deep learning methods have obtained high-quality reconstructed images with complete training samples. However, much training data are required to train the network-trainable parameters, and it is difficult to obtain in many cases. Sidky *et al*. lead to the AAPM challenge named DL-spectral CT. This challenge provides an opportunity for investigators in SCT image reconstruction, using data-driven or iterative techniques, to compete with their colleagues on the accuracy of their methodology for solving the inverse problem associated with spectral CT acquisition from incomplete, noiseless measurements.^a^

This study aims to demonstrate whether deep neural networks can realize high-precision decomposition images of basis materials and establish a foundation for SCT reconstruction algorithms based on big data and deep learning [41]. We propose an iterative network method that fuses SCT physical processes and traditional mathematical algorithms. Although the AAMP challenge provides accurate physical processes (including spectrum and attenuation coefficient information), our method describes the physical process and relevant physical quantities as learnable network parameters, which is more flexible. As with DL-sparse-view CT, high reconstruction accuracy is only possible if the forward model is explicitly incorporated into the solution map, e.g., by an iterative promotion of data-consistency [44]. Here, we also build physical process into precise physical module, which can realize the accurate expression of the positive process in iterative steps, so as to realize the residual transmission correctly. For the noisy case, we add regularization terms, which is more practical. The main contributions of our work are as follows:

- The proposed method is based on the complementary advantages of physical processing, traditional mathematical iteration algorithms (TMIAs), and deep learning. Physical processing realizes the accurate expression of physical phenomena, TMIAs robustly achieve domain transformation, and deep learning can accurately materials decomposition.
- A network joint physical model governs SCT projection data generation. Hence, the network can more accurately express the SCT physical process. Only when the physical process can be accurately and adequately described by one model can the iterative process continuously improve reconstruction accuracy. The physical model of SCT is defined as a module of the network whose x-ray spectra *S*_*k*_(*E*) and attenuation coefficient *θ*(*E*) correspond to the learnable parameters.
- In the iteration process, the input/label data of networks use residual-to-residual to train the network model, rather than image-to-image or imageto-residual. Because places with large errors are observable in the residual image, it is difficult to distinguish them in the image. Moreover, residual-to-residual builds into the residual attention mechanism. Attention operations in residual images make network training easier.
- The network structure is combined with practical problems. The SCT projection data and reconstructed image are domain-related, but even in inconsistent projections, the associated region is relatively small. Therefore, CNN can apply SCT, but the receptive field should not be generous, so as to avoid phenomena such as overfitting and poor robustness.

## 2 Method

### 2.1 JLRM architecture

We formulate SCT reconstruction as a nonlinear optimization problem to solve basis material images 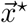 from acquired polychromatic projections 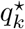:

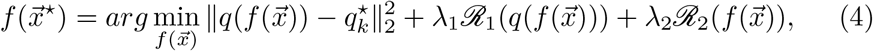

where 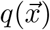 corresponds to the nonlinear SCT physical process, and *ℛ*_1_ and *ℛ*_2_ are network regularization terms in the projection and image domains, respectively, which are used for the case of existing noise or scattering. In this way, *ℛ*_1_ can more accurately describe the physical model for the noise or scattering case. *ℛ*_2_ can effectively suppress noise or scattering and obtain better reconstructed images.

In our experiments, the traditional mathematical iteration algorithm (TMIA) uses the SART algorithm during iteration process and the other TMIA also support. The third step is worth noting. We first calculate the projection residuals for reconstruction, and the results are input to the CNN to obtain the residuals of the base material images. That is, we use the residual-to-residual strategy for network training, which can more effectively improve the image quality.

Generally, the initial value also has a pronounced influence on the convergence speed of the iterative algorithm. Therefore, we incorporate a pre-decomposition module before the iterative process.

Our SCI network includes a pre-decomposition module and an iteration process, as shown in Fig. 2. The pre-decomposition module includes TMIA and CNN steps. For TMIA, we use the E-ART method, which is not sensitive to spectral information, and the quality of reconstructed images is higher than with other analytic algorithms. For the iterative process, the three steps of the incremental method are consistent. As mentioned above, we use a residualto-residual strategy for effective network training. An attention mechanism is incorporated before the residual image input CNN, for easy network training and quick convergence. Below we show pseudocode for the training stage (Algorithm 1) and test stage (Algorithm 2).

**Fig. 2.**
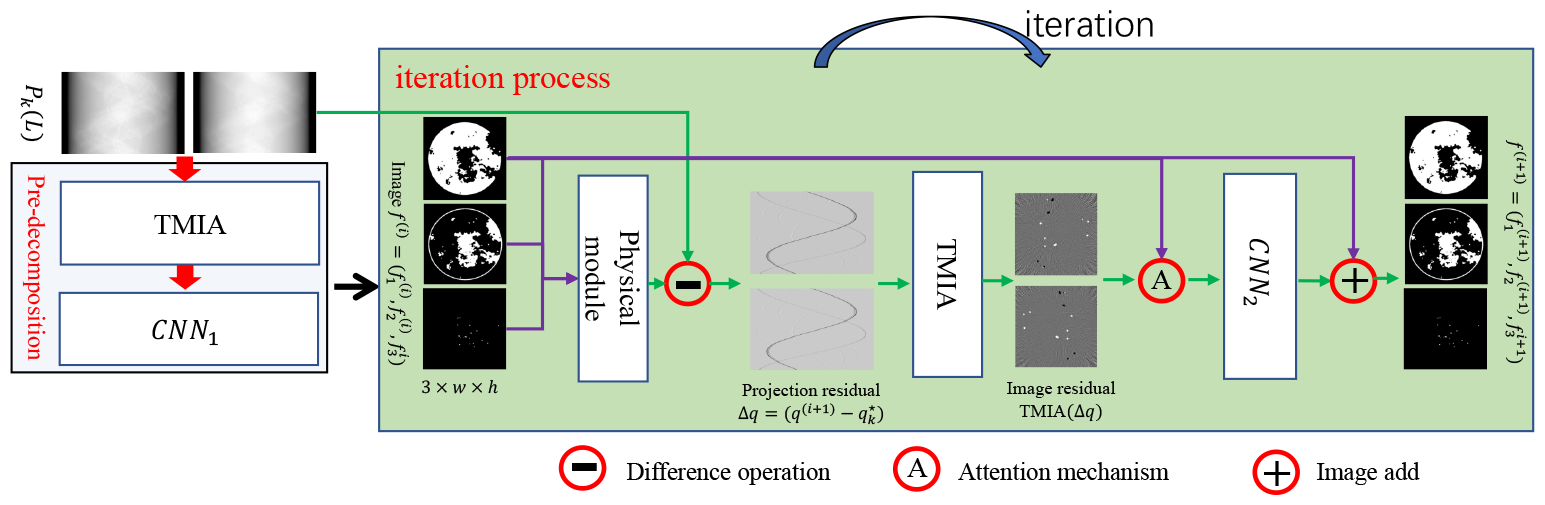
Architecture of proposed method, including pre-decomposition module and iteration process.

#### Algorithm 1

Training stage

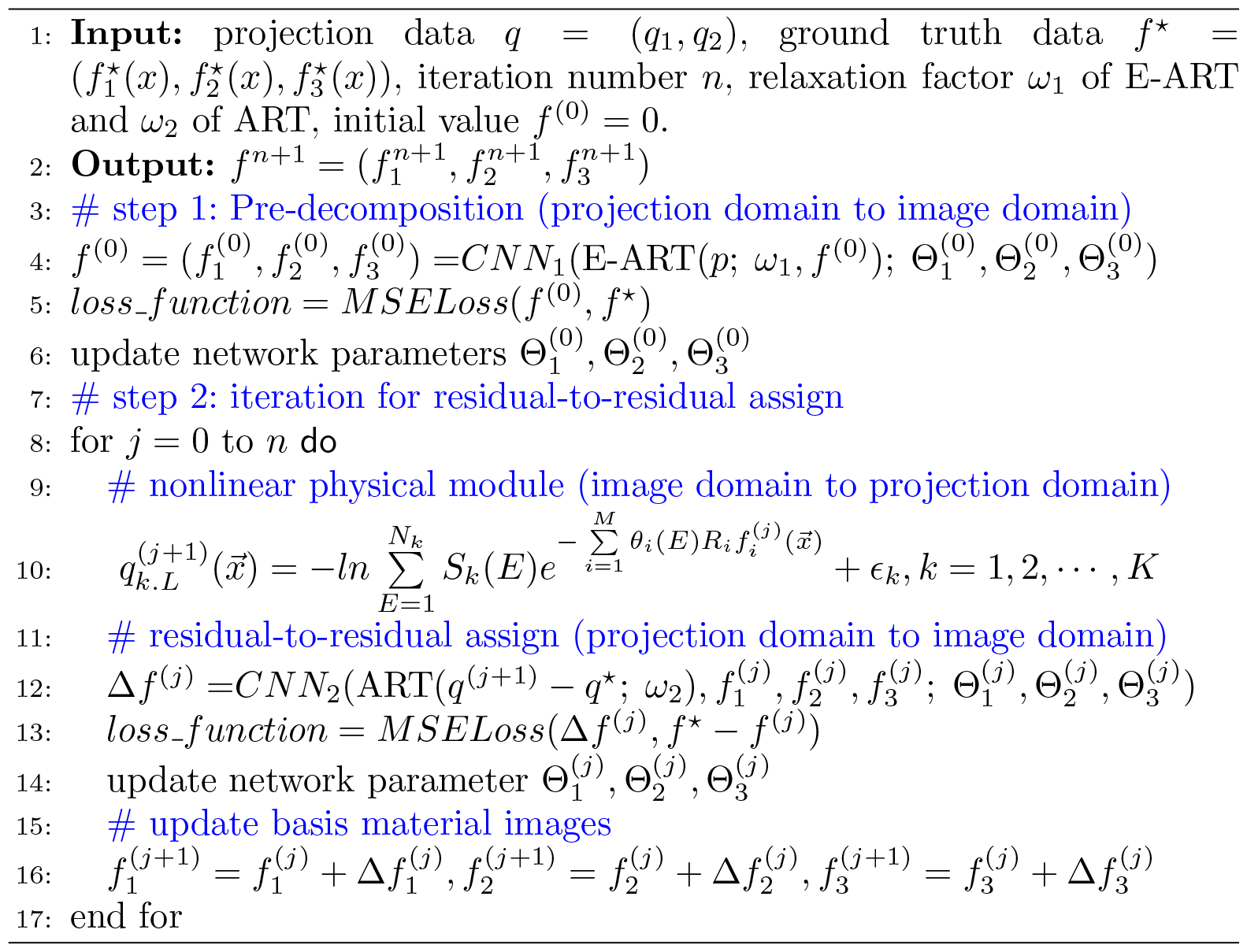

The proposed method is based on the complementary advantages of physical processing, TMIAs, and deep learning. Physical processing realizes the accurate expression of physical phenomena, while TMIAs achieve domain transformation, and deep learning can accurately materials decomposition. The three parts are interdependent and complementary.

#### Algorithm 2

test stage

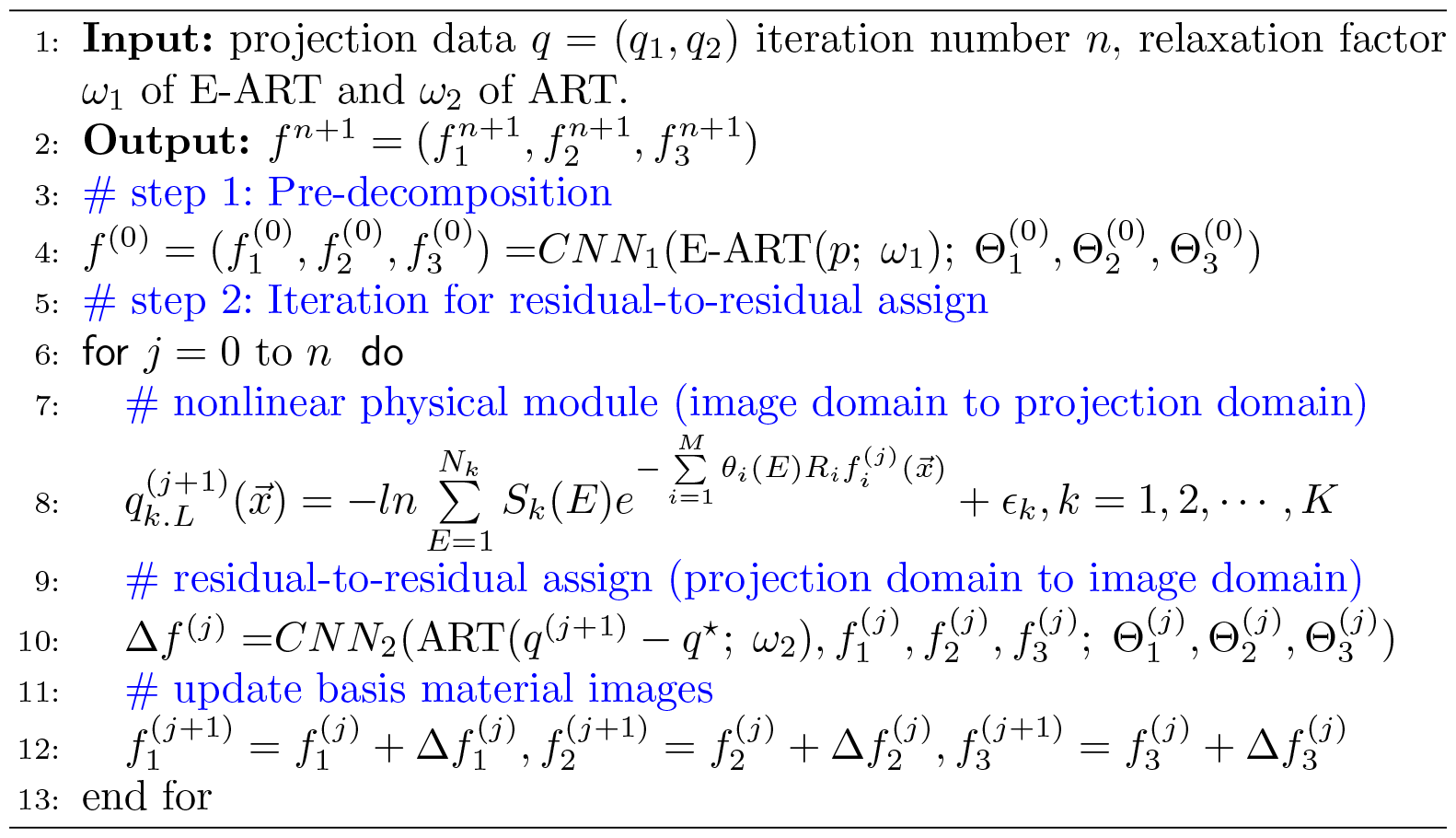

#### 2.2.1 Physical module for nonlinear physical process

Only when the physical process can be accurately described by the model can the iterative reconstruction process continuously improve reconstruction accuracy. Here, to accurately describe the nonlinear physical process of SCT, its positive process is equivalently mapped into a network module. As shown in Fig. 3, Firstly, do a Radon transform for three basis material. This can be achieved by a projection layer [45, 46]. Here, we have overridden the interface which projection and back-projection operators are provide by official. Next, the module can be constructed according to the discrete polychromatic projection formula, where the spectral information and attenuation coefficient can be set as corresponding learning parameters. In fact, both sets of learnable parameters can be achieved by a convolution operator. For the attenuation coefficient, this layer can set K convolution kernels, of size *N*_*k*_ *× M ×* 1 *×* 1 (*out channels × in channels × height × width*). For spectral information, the size of K convolution kernels is 1*× N*_*k*_*×* 1*×* 1. For the noise case, a regularization term network should be added in the physical module to more accurately express the physical process.

**Fig. 3.**
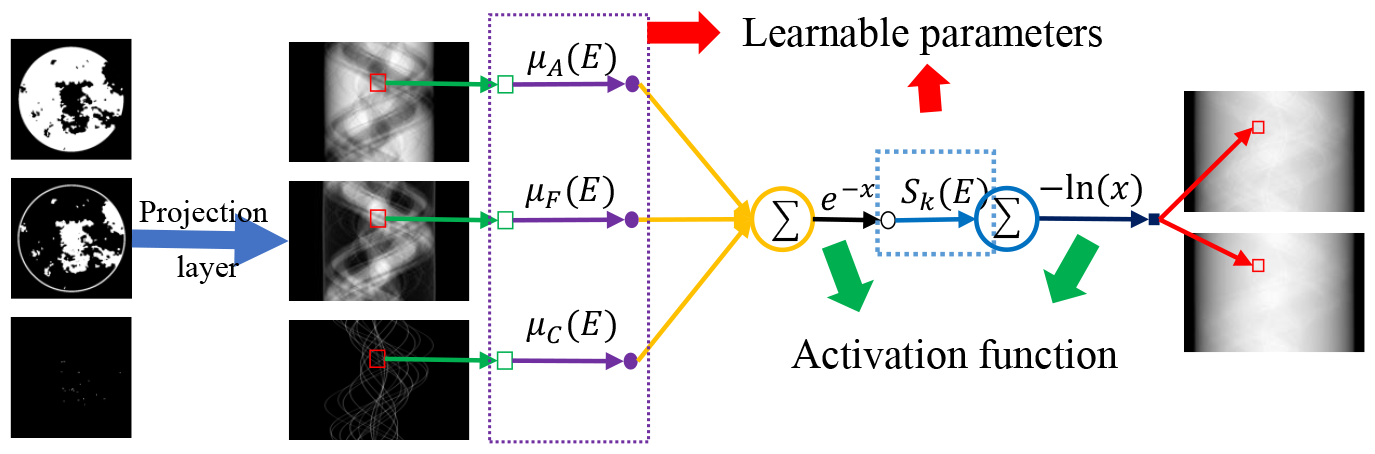
Physical process equivalent mapping physical module.

#### 2.1.2 CNN network structure design

The network structure is also critical to the reconstruction result, because our network is trained block-to-block. Once overfitting occurs, the results of subsequent iterations are disastrous. The CNNs of pre-decomposition and iterative processes are basically similar, with structures as displayed in Fig. 4. The biggest difference between them is the channel setting of input data, in which the channel of *CNN*_1_ is set to 2, while that of *CNN*_2_ is 6. The CNN contains eight blocks, each with three convolution layers, and the kernel sizes are 3×3, 1×1, and 3×3. It is worth noting that network operations such as downsampling, upsampling, and transpose convolution, which make the receptive field large, cannot be used. Although the projection is inconsistent and the pixels of basis material images are domain-related, the regional correlation caused by the inconsistency is very small, and increasing the receptive field will aggravate overfitting, which is fatal for later iterations to obtain more accurate decomposition results.

**Fig. 4.**
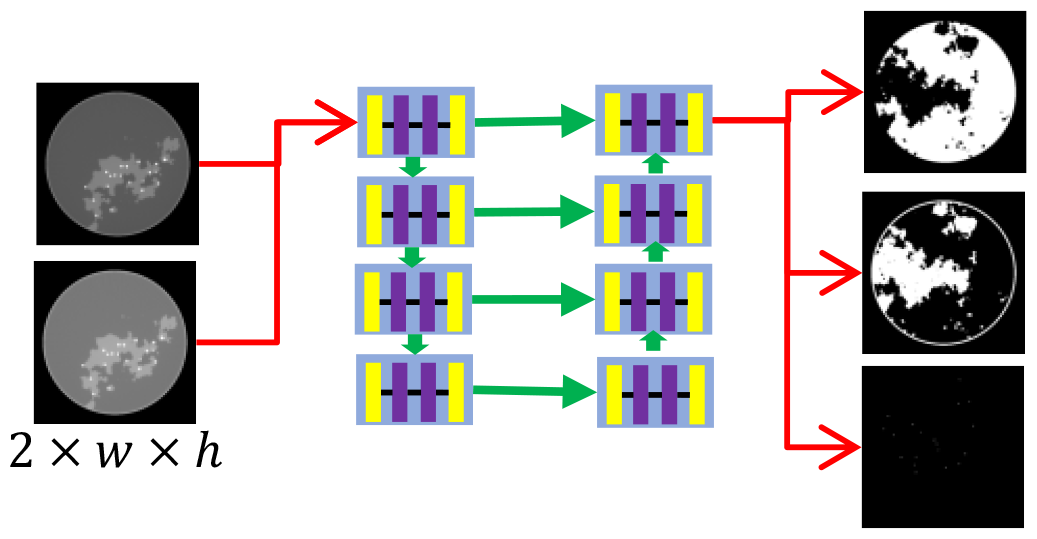
CNN network structure with eight blocks, each with three convolution layers.

#### 2.1.3 Residual-to-residual strategy

The residual-to-residual strategy proposed based on experiments observations which can effectively improve the accuracy of reconstruction. As shown in Fig. 5, regions with large errors can be directly located in the residual image, where it is difficult to find in reconstructed images. Hence, the image-to-residual strategy may bring new errors, with many uncertainties to improve the quality of reconstructed images. In other words, image-to-residual is unstable for improving image quality.

**Fig. 5.**
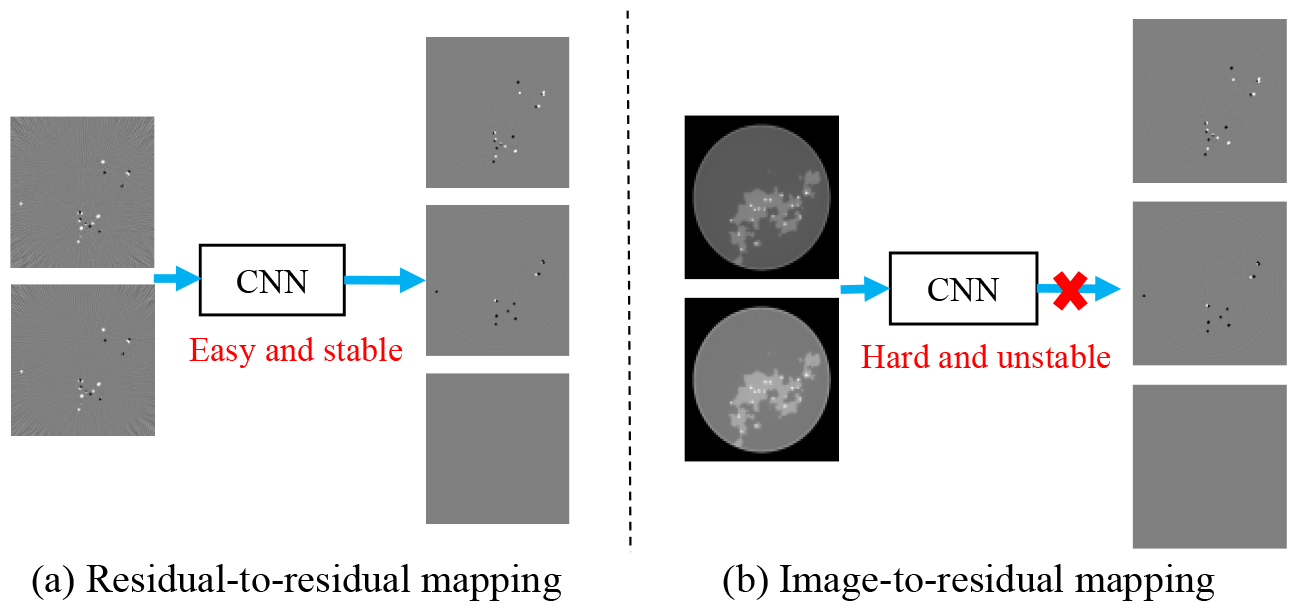
Residual-to-residual vs. image-to-residual strategy. Residual-to-residual strategy makes it easier to locate where there are large errors.

The ReLU activation function is often used in the network, which makes the images obtained by the network nonnegative. The residual image does not possess this property. To use the nonlinear ReLU activation function, we decompose the residual image *dx* into the form

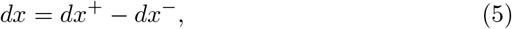

where *dx*^+^ and *dx*^*−*^ are, respectively, the positive and negative parts of the residual image. Thus, the network output becomes an image with two nonnegative channels.

#### 2.1.4 Residual attention mechanism

Generally speaking, in reconstructed residual images, even if the decomposition is correct, there are still some errors, and this phenomenon occurs at the edges of basis material images. This can be overcome by incorporating an attention mechanism 47–49] to emphasize the edges, which can make the network training more simple and efficient. Attention includes both channel and spatial attention. In our case, we need to emphasize edges that are spatial features, so spatial attention is employed. Given a tensor of intermediate feature map *η ∈ R*^*c×h×w*^, spatial attention generates a spatial attention map *A ∈ R*^1*×h×w*^ by utilizing the inter-spatial relationships of features. Then the output of the attention mechanism, 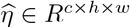, can be expressed as

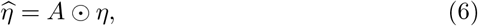

where ⊙ denotes element-wise multiplication and *c* is the channels of input data. During multiplication, the attention values are broadcast (copied) to all feature maps along the channel dimension.

The spatial attention focusing on “where” is the most informative part. Considering that there are three materials, we implement the attention mechanism in three directions.

As shown in Fig. 6, our attention mechanism consists of three pipelines to construct three spatial attention maps. Given a two-dimensional feature map

**Fig. 6.**
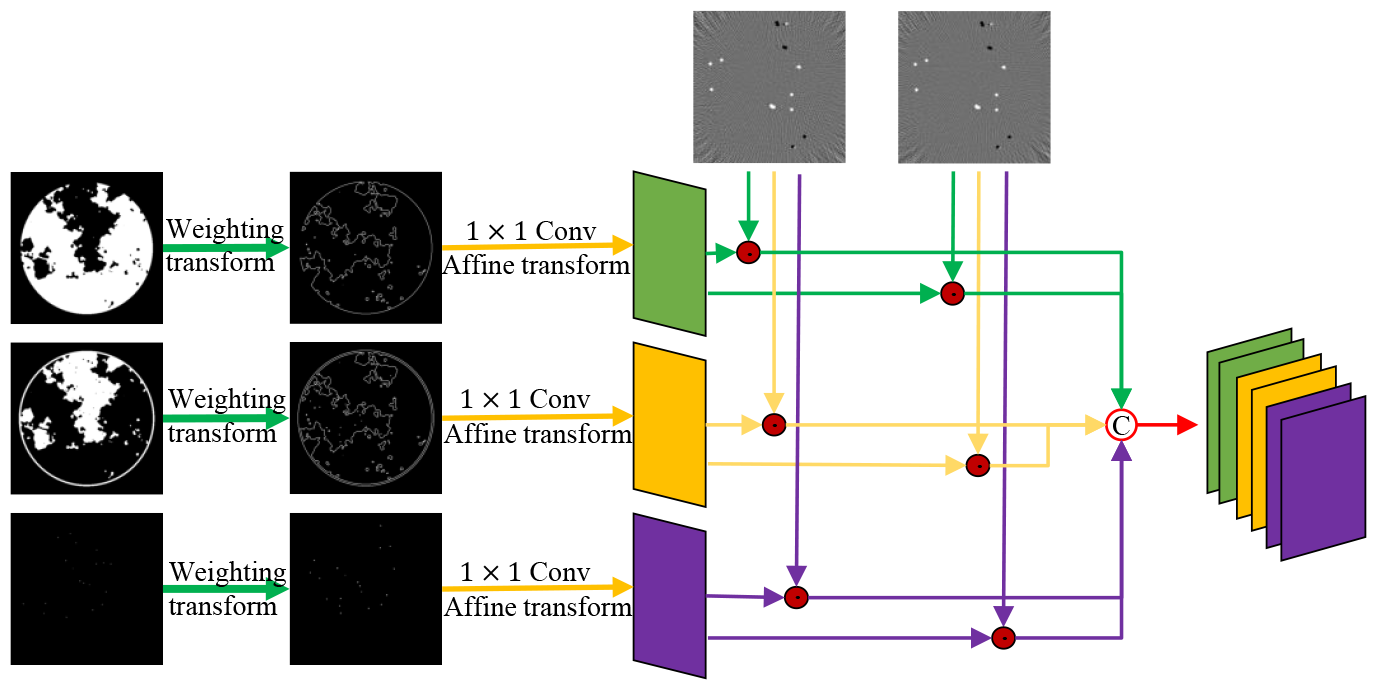
Residual attention mechanism.

*f ∈ R*^*h×w*^, which is obtained by applying the weighting transform on three basis material images, let us define

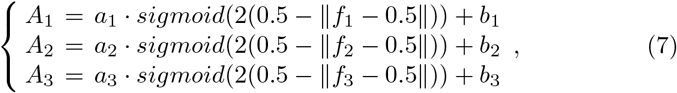

where *a*_1_, *b*_1_ and *a*_2_, *b*_2_ represent the weight parameters and bias in the 1×1 convolutional layer, and *sigmoid* denotes the sigmoid activation function. Here, we set the initial values as *a*_*i*_ = 1, *i* = 1, 2, 3, and *b*_*i*_ = 0, *i* = 1, 2, 3. To apply the attention mechanism, each residual image element-wise multiply spatial attention maps and obtained feature maps would have been re-weighted, which is helpful to accelerate the convergence of the network.

### 2.2 Network training

The proposed method was implemented using Python under the PyTorch framework. Each module was trained module-by-module, and the network was optimized using the Adam optimization method with a min-batch of nine image patches for each iteration. The objective function for each module used an MSE loss function,

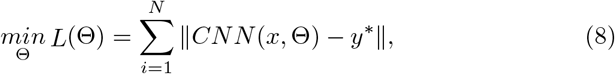

where *x, y*^*∗*^ are the input/label data, and Θ denotes the network learnable parameters. The complete training samples were randomly shuffled, and as an empirical stopping criterion of each module, the number of training epochs was set to 1000. Comparative experiments were performed on a PC (AMD EPYC-Rome processor, 128 GB RAM). Training was executed on a single graphic processing Nvidia RTX A6000 with 48 GB of video memory.

## 3 Results

Experiments were carried out to demonstrate the behavior of the proposed method, including architecture validation, for example, residual analysis and the performance of JLRM in noise suppression, reconstruction method, CNN network structure design and residual-to-residual strategy.

### 3.1 DL-spectral CT setup of AAPM challenge

This challenge focuses on spectral CT, where the object is scanned by X-rays with two spectra, i.e., *K* = 2, without noise. This challenge model an ideal form of fast kVp-switching where the X-ray tube voltage alternates between two settings for consecutive projections. For each kVp setting, the beam spectrum is broad and the polychromatic nature of the X-ray beam makes quantitative imaging in CT quite challenging when only one kVp setting is used. Scanned objects are simulated for a breast model [50] that contains three tissue types (*M* = 3): adipose, fibroglandular, and calcification. Truth images and simulated data are provided for 1000 cases so that participants can decide to use either a data-driven or optimization-based technique. The SCT imaging process can be seen in Fig. 7, which shows the spectral information used in the imaging process and the linear attenuation coefficient of the material. It can be observed that the linear attenuation coefficients of the two materials are very close. This is a tricky point, as discussed in the experimental results. To be fair, the test dataset adopts 10 additional cases, which are provided in a “starting kit” with ground-truth images. The fan-beam geometric parameters of the SCT system are listed in Table 1. The DL-spectral-CT challenge seeks the image reconstruction algorithm that provides the most accurate reconstruction of the adipose, fibroglandular, and calcification spatial maps from SCT projection data. Details about the challenge setup and results can be found on the official website.^b^

**Table 1.** System and geometric scanning parameters of AAPM DL-spectral CT challenge dataset.

|  |  |
| --- | --- |
| X-ray source to rotation center (mm) | 500 |
| X-ray source to detector distance (mm) | 1000 |
| Scanning angle (deg) | $[0^\circ, 360^\circ]$ |
| Scanning angular interval (deg) | 1.40625 |
| Number of detector units | 1024 |
| Detector unit size (mm) | 0.3574 |
| Reconstructed image size (pixel) | $512 \times 512$ |

**Fig. 7.**
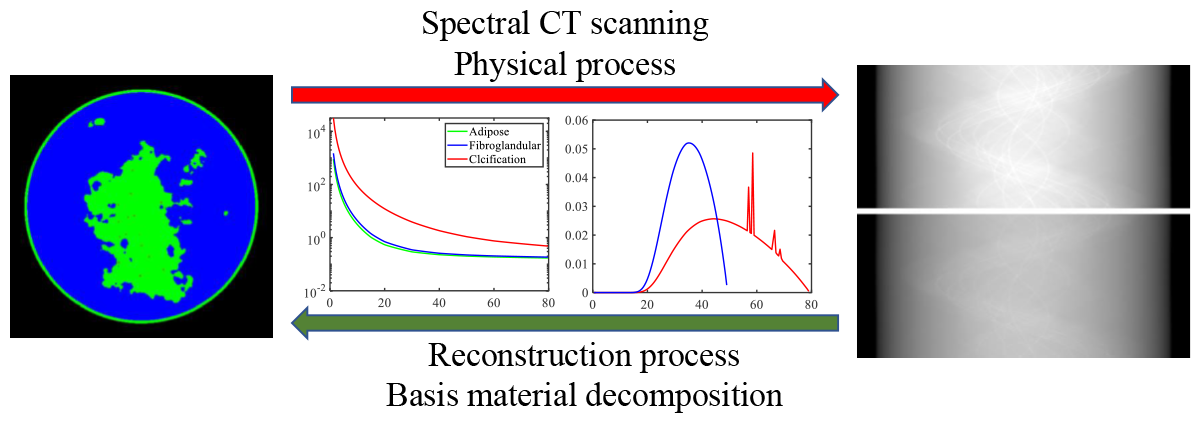
SCT imaging, where forward process is data collection, which is also a physical process. Inverse process is reconstruction, i.e., basis material decomposition.

### 3.2 Performance and analysis of JLRM

We demonstrate that JLRM can be trained to accurately reconstruct basis material images from collected projection data obtained from the DL-spectral CT of the AAPM challenge. As described in section 2, the JLRM method includes a pre-decomposition module and two iteration of the iteration process. Fig. 8 shows the basis material images of these three modules. It is difficult to visually distinguish the image quality reconstructed by each module. The last row indicates the tissue label map of the three materials, from which we can clearly see the proportion of each material in each pixel. Fig. 9 shows the different images. The accuracy of the proposed method can be further improved, and the sequence module can continuously improve the accuracy of reconstruction based on comparison with the differential images of each module. Even in the display window with almost mechanical accuracy, the difference images of the final result (2 iterations) are difficult to indicate a large error. This can verify that our method can achieve near exact reconstruction. Table 2 shows the average RMSE for quantitative evaluation, which further indicates that our method can obtain accurate reconstruction. The accuracy of the results finally submitted in the AAPM challenge is 6.8×10^*−*7^. Report to the second place accuracy of 6.21 ×10^*−*6^, our reconstruction accuracy is higher one order of magnitude.

**Table 2.** Average RMSE for quantitative evaluation.

|  |  | Pre-decomposition | 1 iteration | 2 iterations |
| --- | --- | --- | --- | --- |
| adipose | training set | $2.53 \times 10^{-3}$ | $9.24 \times 10^{-5}$ | $6.79 \times 10^{-6}$ |
| | test set | $2.87 \times 10^{-3}$ | $7.09 \times 10^{-5}$ | $7.54 \times 10^{-7}$ |
| fibroglandular | training set | $2.68 \times 10^{-3}$ | $1.20 \times 10^{-4}$ | $5.54 \times 10^{-6}$ |
| | test set | $3.06 \times 10^{-3}$ | $9.74 \times 10^{-5}$ | $6.67 \times 10^{-7}$ |
| calcification | training set | $3.30 \times 10^{-7}$ | $6.60 \times 10^{-7}$ | $3.00 \times 10^{-7}$ |
| | test set | $6.40 \times 10^{-7}$ | $6.30 \times 10^{-7}$ | $6.60 \times 10^{-7}$ |

**Fig. 8.**
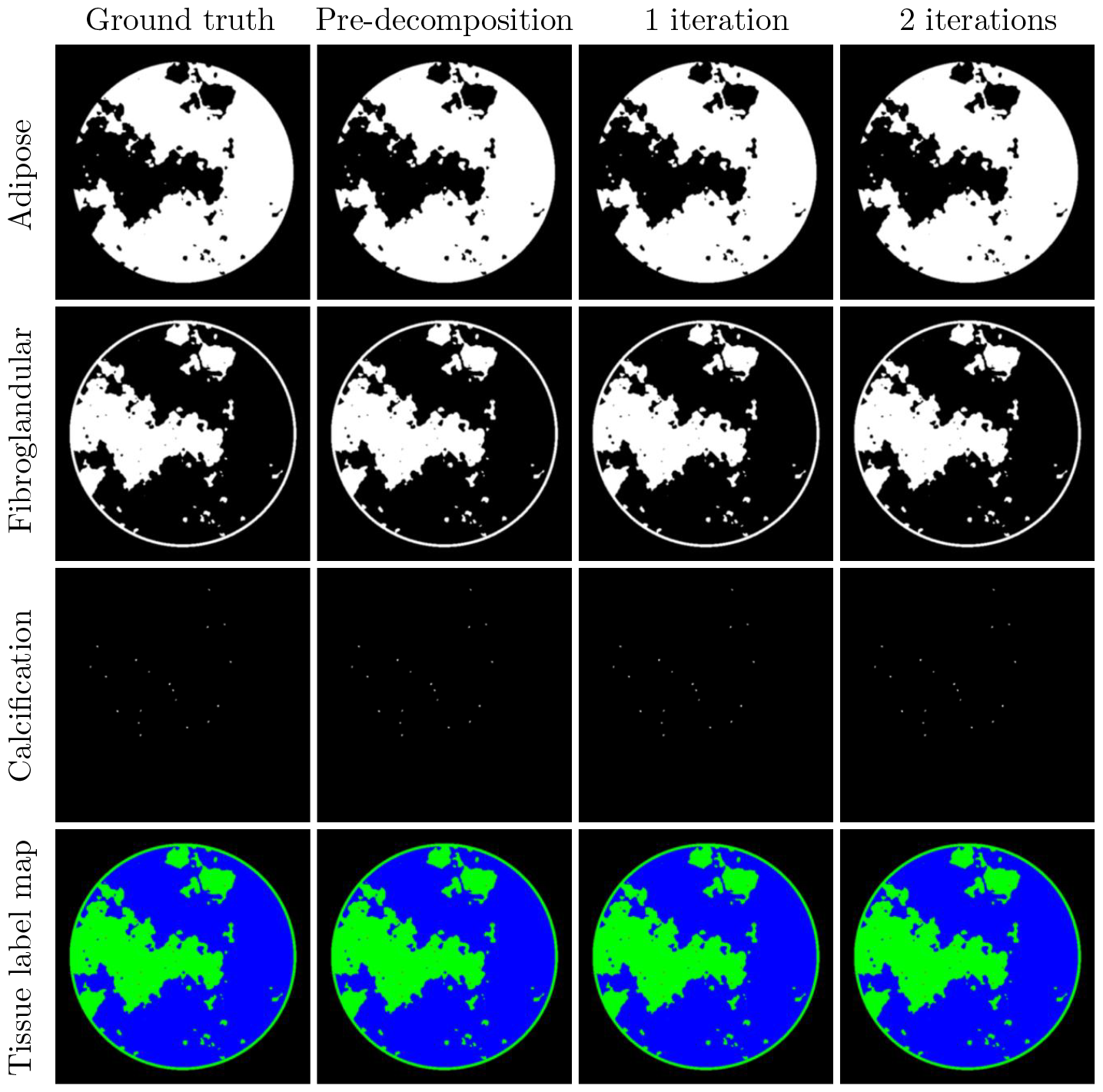
Left to right: ground truth, results of pre-decomposition block, iteration process with 1 and 2 iterations. Display windows: [0.15, 0.35] for adipose material, [0.3, 0.7] for fibroglandular material, [0.1, 0.5] for calcification material. Last row: tissue label map with adipose (blue), fibroglandular (green), calcification (red).

**Fig. 9.**
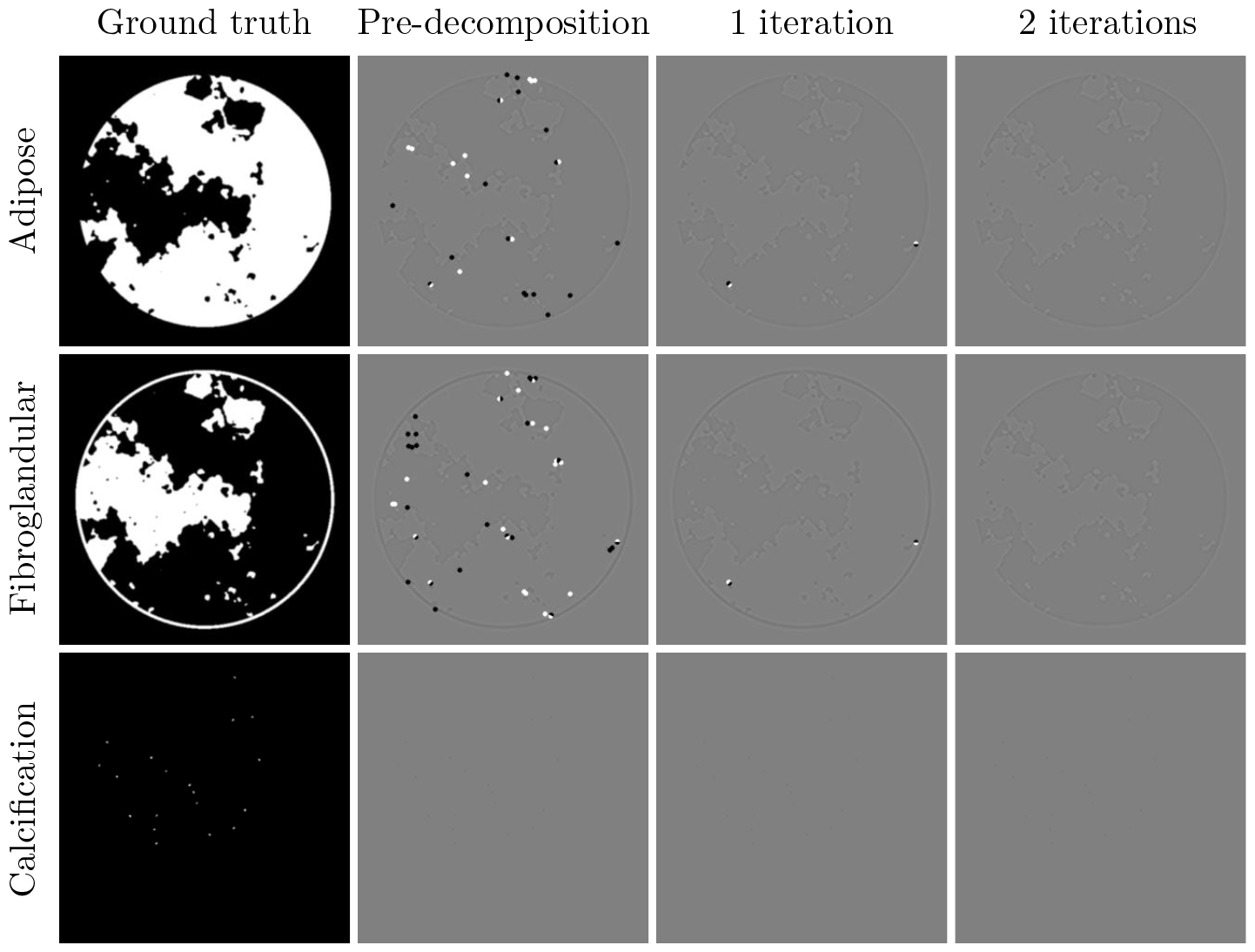
Left to right: ground truth, difference images of pre-decomposition block, iteration process with 1 and 2 iterations. Display windows of difference images are [-0.00005, 0.00005].

To better understand the behavior of each module of the proposed method, Fig. 10 shows the loss function update curve of each material of each module. In the pre-decomposition module, adipose and fibroglandular material cannot reach the mechanical accuracy, but more cases of calcification material can reach the mechanical accuracy. In the iteration process, the accuracy can be well improved using the residual-to-residual strategy.

**Fig. 10.**
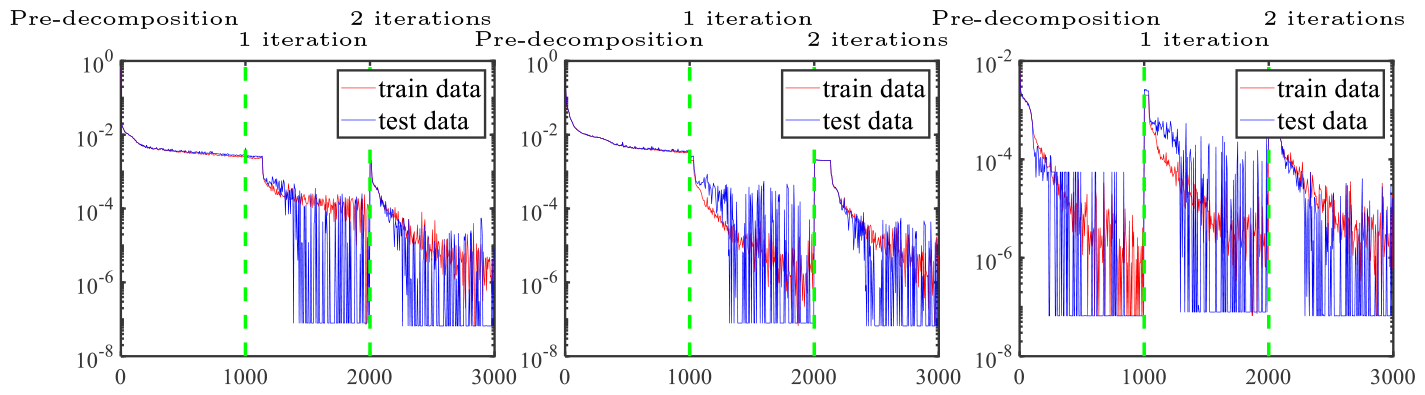
Update curves of three material loss functions. Left to right: adipose, fibroglandular, and calcification material.

### 3.3 Residual analysis

Because the projection data of the AAPM challenge are noise-free, a crucial way of a proper solver for this DL spectral-CT problem is evaluating this difference images 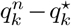, which should be as low as possible. We analyze this aspect in Fig. 11. For better analysis and observation, we reconstructed the residual images from the residual of projection. Fig. 12 displays the reconstructed residual images. We note that the residual images in the image domain also showed a decreasing trend. The results of the final iteration showed that each pixel of the residual was all within the mechanical accuracy.

**Fig. 11.**
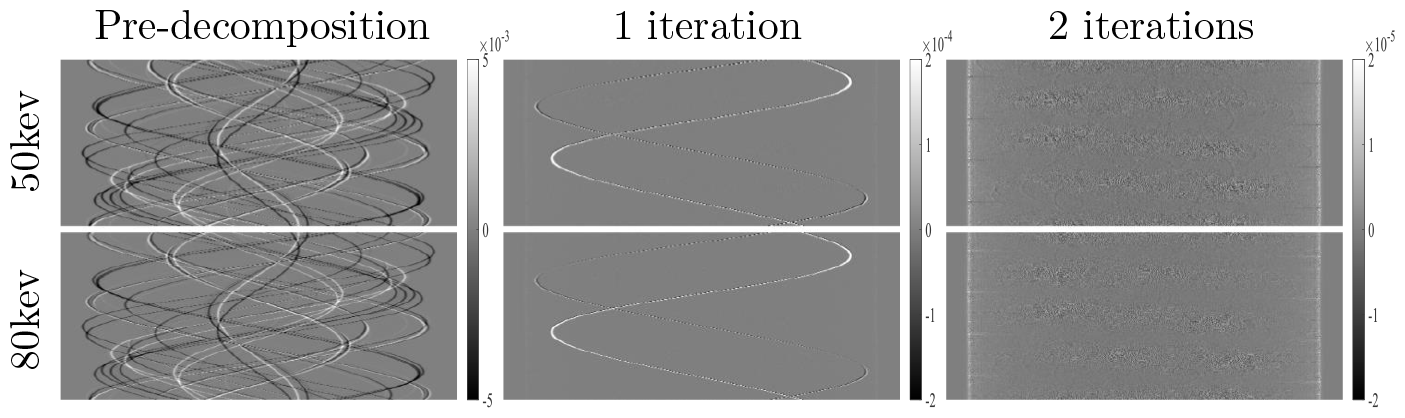
Errors of projection domain for each module.

**Fig. 12.**
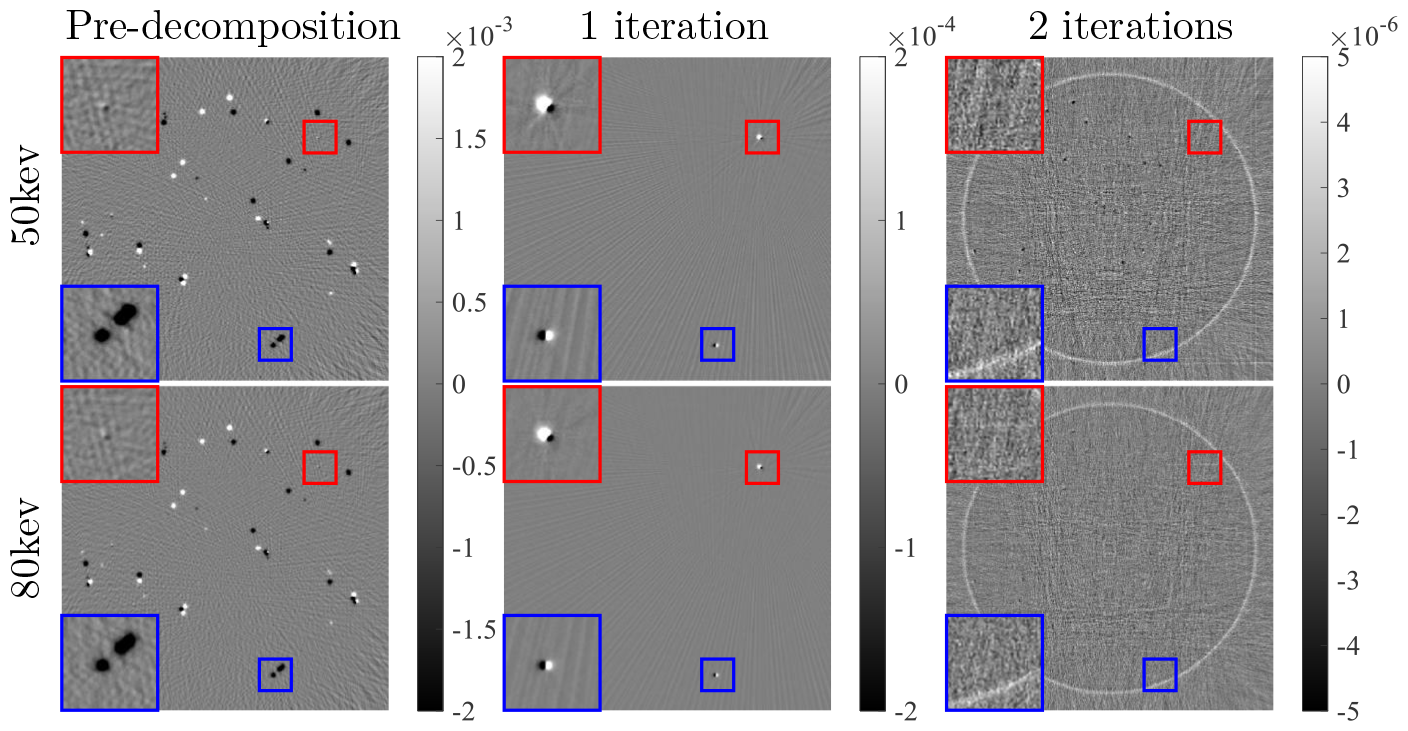
Errors of image domain for each module. Colored boxes mark trend of updates during iterative process; corresponding zoomed-in views are displayed in left corner.

Some pixels with small residuals, marked by red boxes in Fig. 12, cannot be updated effectively in the first iteration, and are shown to be more obvious in the residual image after one iteration, which facilitates further performance improvement. Some regions with large errors can continuously shrink during the iteration process, although updates still have larger errors, as marked by blue boxes.

### 3.4 Performance of JLRM in noise suppression

To ensure universality, we study the noise data. The statistics of x-ray measurements are often described using a Poisson distribution to model the incident intensity,

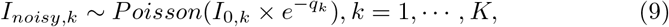

where *I*_*noisy,k*_ and *q*_*k*_ denote received intensity of detectors and noise-free projection data, respectively. The noisy projection data 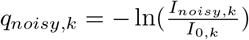. Table 3 lists the accuracy of the proposed method under different noise levels. As shown in previous experimental results, the phenomenon of adipose and fibroglandular materials is also consistent, mainly because the attenuation coefficients of the two materials are very similar. Hence, the two materials are easily decomposed incorrectly, and their performance evaluation indices are basically consistent. The attenuation coefficient of calcification material is large, and its size is very small; hence, calcification material can still have mechanical accuracy with high noise levels (*I*_0_ = 1*×*10^5^) and can do so quickly when the noise level is relatively low (*I*_0_ = 1 *×* 10^6^).

**Table 3.** Average RMSE of dataset with different noise levels for quantitative evaluation.

| | Noise level | $1 \times 10^5$ | $5 \times 10^5$ | $1 \times 10^6$ |
| --- | --- | --- | --- | --- |
| adipose | training set | $7.84 \times 10^{-2}$ | $2.87 \times 10^{-2}$ | $6.58 \times 10^{-3}$ |
| | test set | $9.97 \times 10^{-2}$ | $4.45 \times 10^{-2}$ | $7.54 \times 10^{-3}$ |
| fibroglandular | training set | $7.23 \times 10^{-2}$ | $3.24 \times 10^{-2}$ | $6.69 \times 10^{-3}$ |
| | test set | $9.17 \times 10^{-2}$ | $4.68 \times 10^{-2}$ | $8.20 \times 10^{-3}$ |
| calcification | training set | $7.07 \times 10^{-5}$ | $3.54 \times 10^{-6}$ | $2.72 \times 10^{-6}$ |
| | test set | $8.92 \times 10^{-5}$ | $5.47 \times 10^{-6}$ | $1.03 \times 10^{-6}$ |

To further understand the effect of noise on the decomposition results of adipose and fibroglandular materials, we analyze a set of results obtained from noise-free pre-decomposition. As in the residual analysis of section 3.3, the obtained projection domain residuals are reconstructed to obtain the results shown in Fig. 13, where the projection residuals are obtained by performing the physical process with the pre-decomposition results minus the noisy projections, i.e., 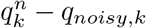. Fig. 13 shows that it is too difficult to reflect some decomposition errors in the image when the noise is relatively large, such as those marked in the red box (zoomed-in view is shown in upper-right corner). However, in some locations where the error is smaller, the noise is small but also difficult to decompose, while the noise-free case can be obviously found, such as in the blue box (zoomed-in view shown in upper-left corner).

**Fig. 13.**
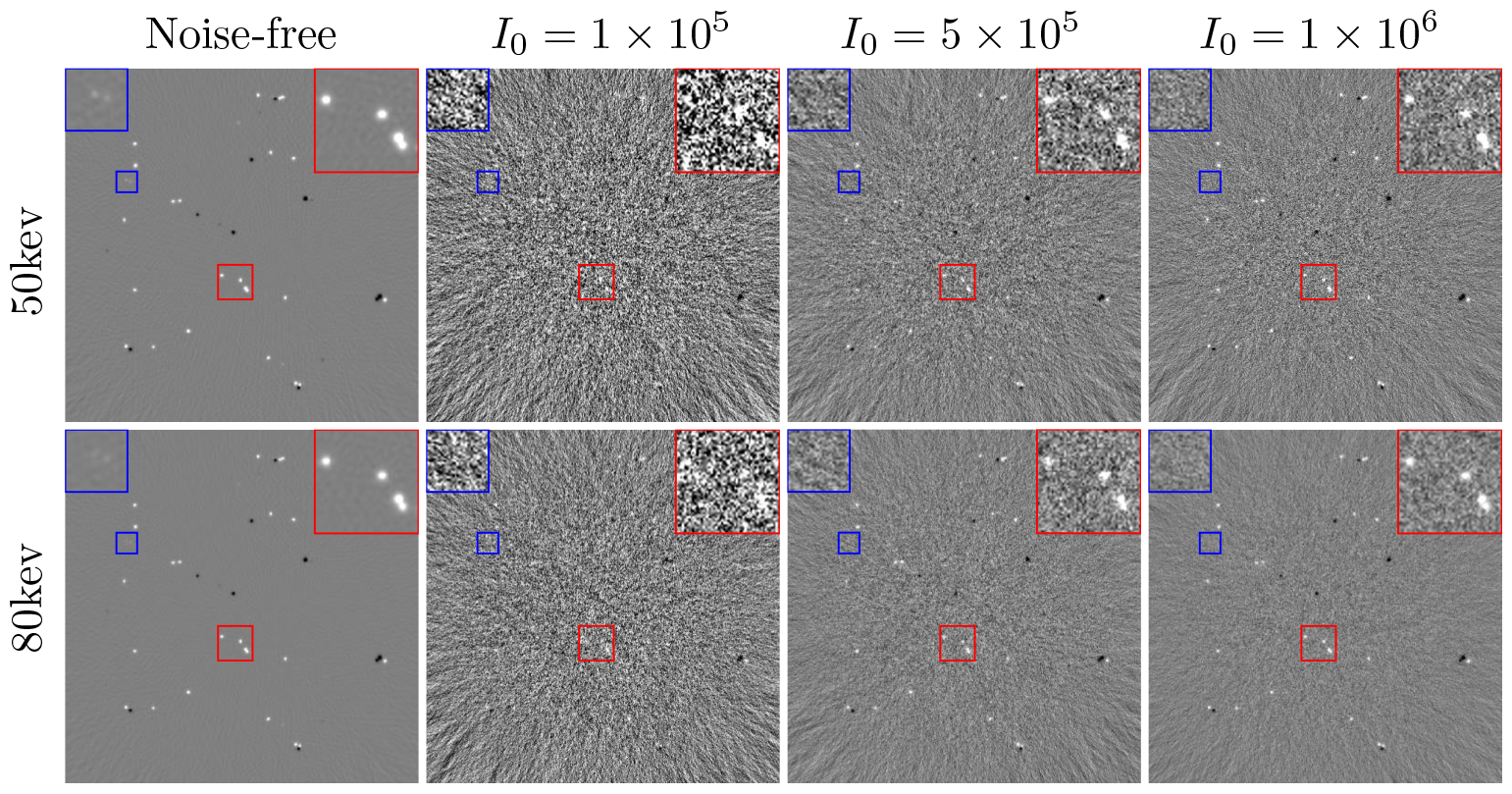
Residual images with different noise levels. Colored boxes mark details of images under different noise levels; corresponding zoomed-in views are displayed in upper corners.

### 3.5 Receptive field: small or large?

As argued in CNN network structure design of section 2 increasing the receptive field will aggravate the overfitting phenomenon, which is fatal for the later iteration to obtain more accurate decomposition results. Fig. 14 shows the loss function curve update of the pre-decomposition module. During the training process of the proposed CNN, the changes of the training and test sets are essentially the same with the iterative update. For UNet, using pooling and upsampling layers causes the network to have a large receptive field. However, the experimental results show that adipose and fibroglandular materials display an obvious overfitting phenomenon after a period of training.

**Fig. 14.**
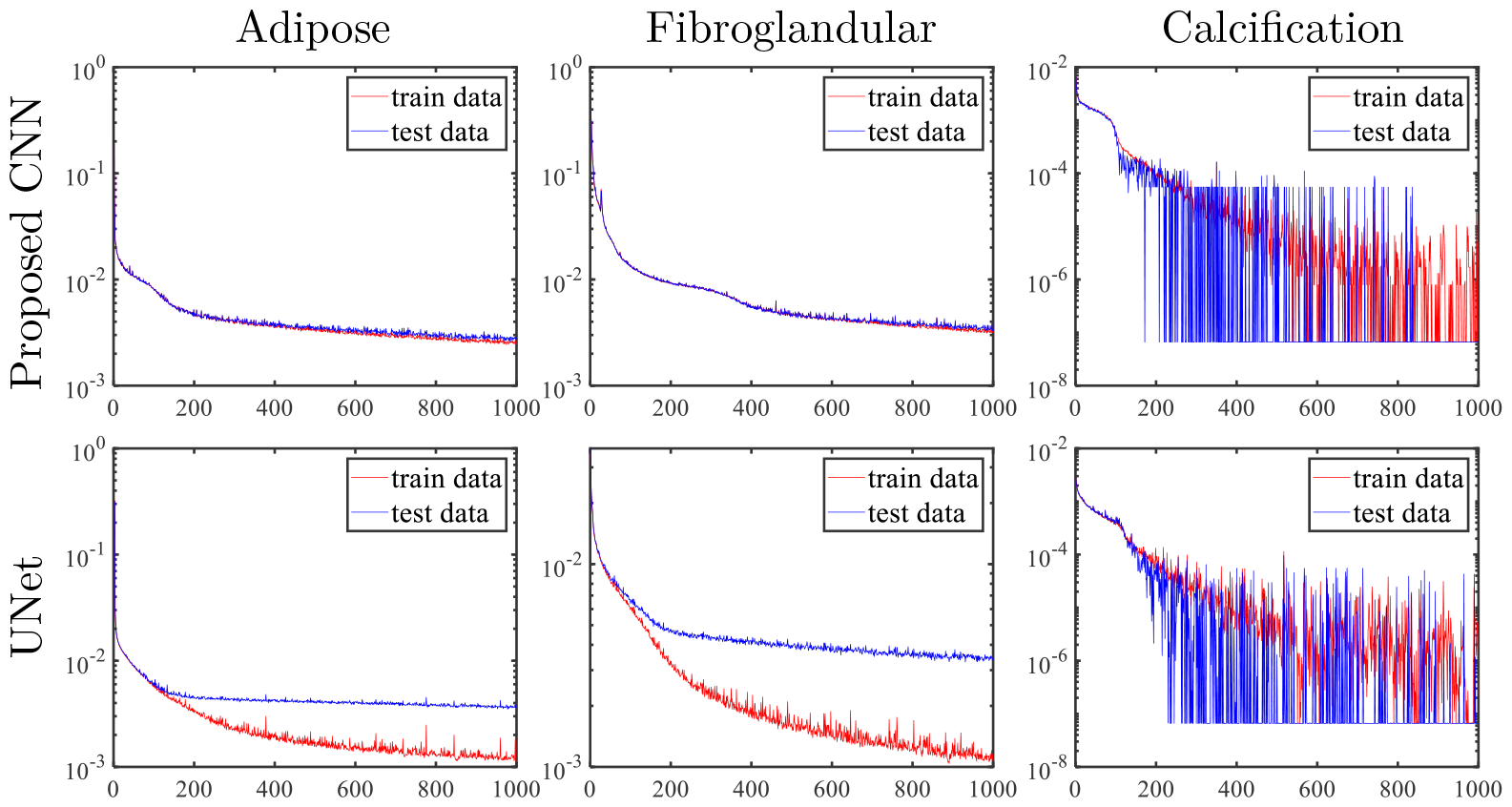
Update curves of three material loss functions for pre-decomposition module with networks with different receptive fields.

Fig. 15 shows the performance of overfitting in the reconstructed image of adipose material, where fibroglandular material has the same phenomenon. The training set has higher accuracy for UNet, and the error in the difference image is quite small, while the test set has a significantly larger error.

**Fig. 15.**
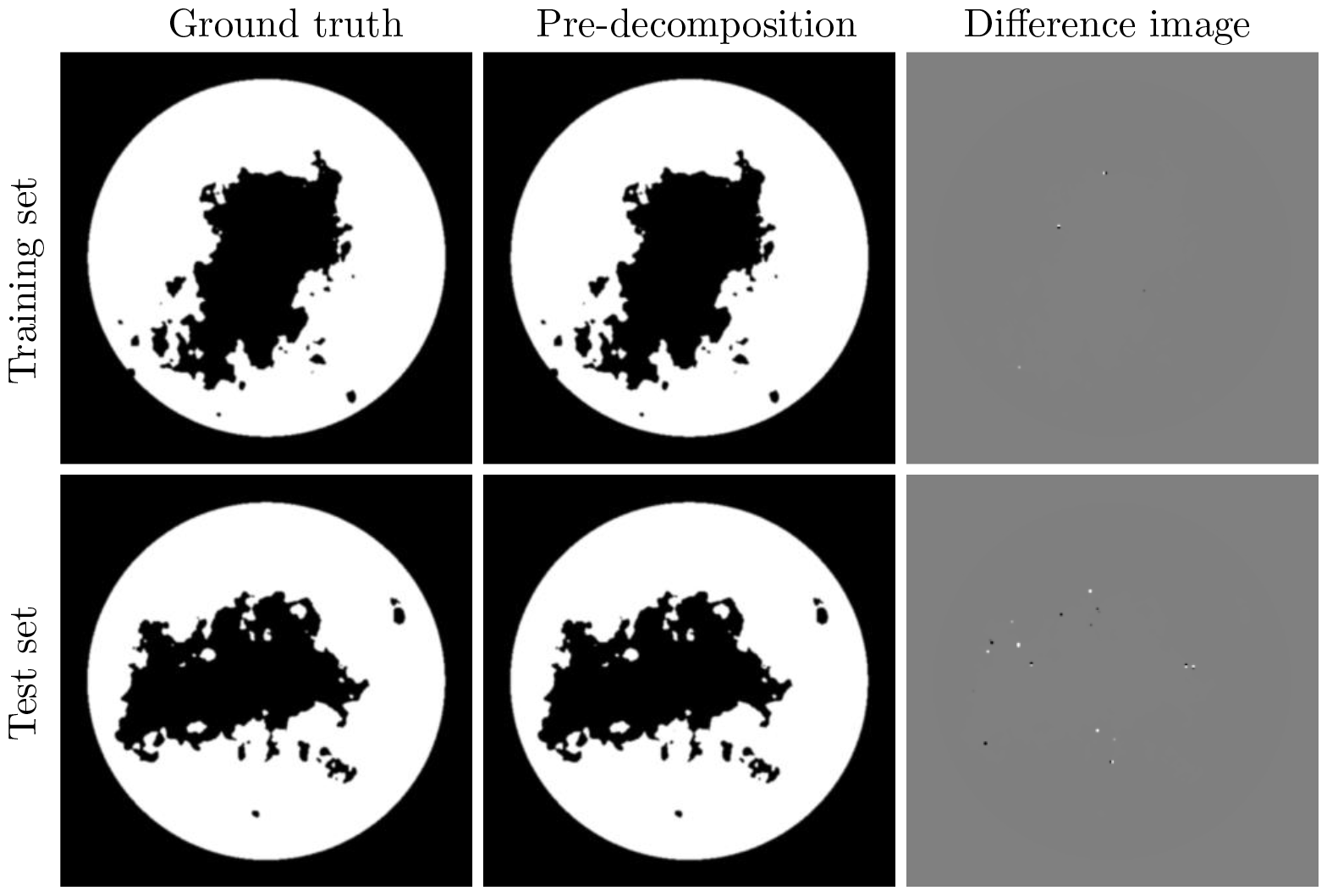
Results of reconstruction of training and test sets using a larger receptive field. Display windows: [0,1] for material images, and [-0.05,0.05] for difference images.

### 3.6 Residual-to-residual vs. image-to-residual

We research the mapping strategy. Fig. 16 displays the partial results of two iterations of the image-to-residual and residual-to-residual mapping strategies for adipose and fibroglandular materials. The image-to-residual mapping strategy may bring new reconstruction errors, so that the accuracy of the final reconstruction cannot be really improved, and there exists the risk of instability. Due to the places with large errors are difficult to distinguish them in the reconstruction images. The residual-to-residual strategy can effectively improve the accuracy compared with the image-to-residual strategy. The high-precision reconstruction results can be attributed to the following: 1) It can calculate the residual in combination with the physical module; 2) The attention mechanism is used to reduce the complexity of network decomposition; 3) The residual-to-residual strategy avoids large numbers eating decimals.

**Fig. 16.**
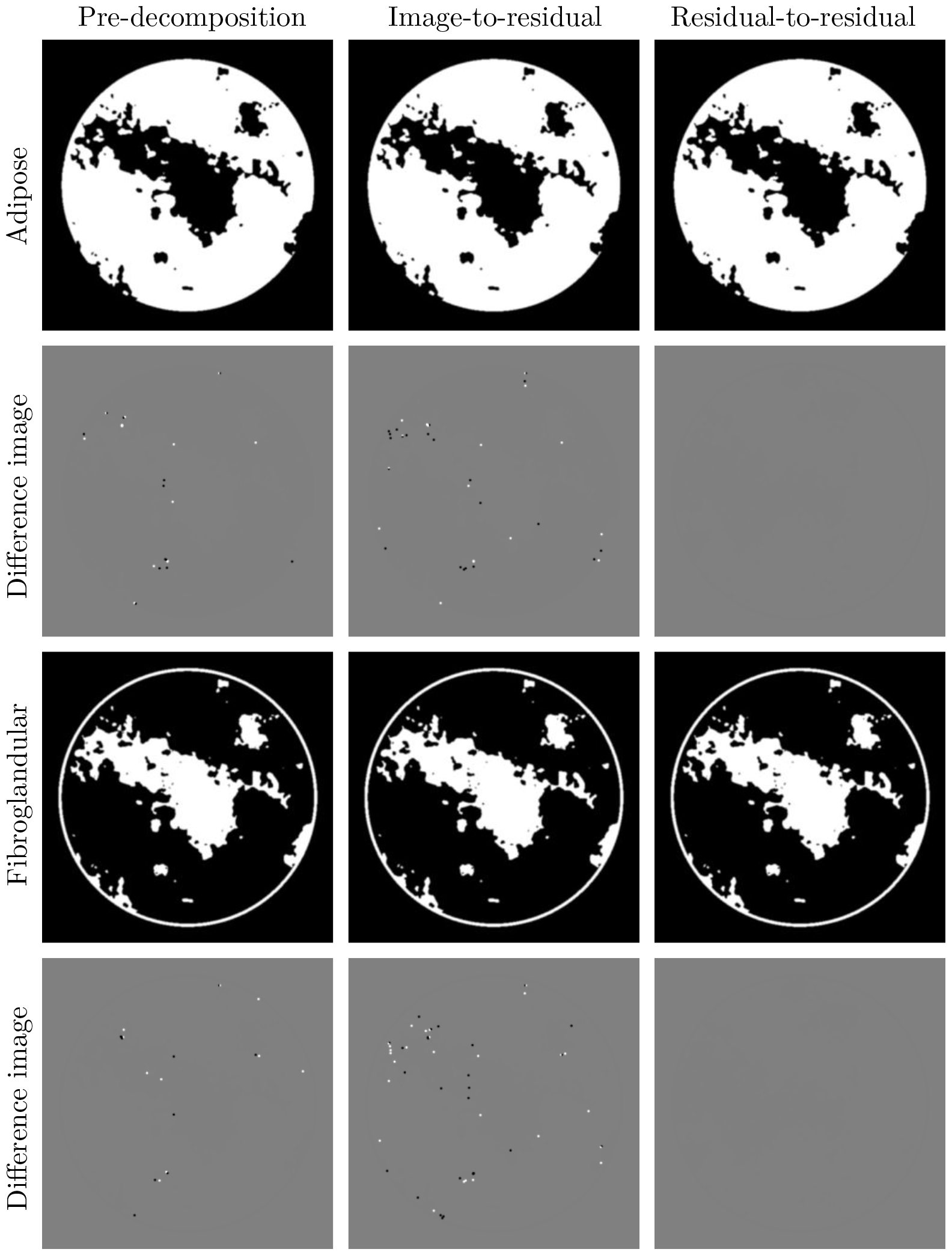
Results of residual-to-residual and image-to-residual mapping strategies. Display windows: [0,1] for material images and [-0.05,0.05] for different images.

## 4 Discussion

To solve the problem of high-precision base material decomposition in spectral CT, this paper proposes an algorithm fused on a physical model, traditional mathematical iterative algorithm, and deep learning. Each part has clear functions and is built based on given problems and actual situations. The physical module, which is embedded and expressed as a network structure, can accurately describe the physical process of SCT imaging. The mathematical iterative algorithm realizes information transfer between domains. These three modules complement each other. We introduce some network skills or strategies in some modules, which are observations related to SCT problems, and can improve the reconstruction results. Examples are a residual attention mechanism and the study of the receptive field.

To further illustrate the effectiveness of our method, we study it when the data are noisy.

The JLRM algorithm is based on deep learning, which mixes mathematics and physics. It can obtain satisfactory decomposition results due to a large number of training datasets. In practice, it is difficult to obtain a large volume of SCT projection data, and it requires matching label data (basis material images). The current energy spectrum is based on the data generated under a single specific energy spectrum. However, the spectral curves of the actual scanning system under the same energy are not necessarily consistent.

## Data Availability

All data produced are available online at https://dl-sparse-view-ct-challenge.eastus.cloudapp.azure.com/competitions/3#learn_the_details

## Acknowledgments

This work was supported by the National Natural Science Foundation of China (No. 61827809) and the National Key Research and Development Program of China (No. 2020YFA0712200). The authors are grateful to the Beijing Higher Institution Engineering Research Center of Testing and Imaging and the Beijing Advanced Innovation Center for Imaging Technology for funding this research work.

## Footnotes

a https://dl-sparse-view-ct-challenge.eastus.cloudapp.azure.com/competitions/3

b https://dl-sparse-view-ct-challenge.eastus.cloudapp.azure.com/competitions/3#learn_the_details

## Notes

### Competing Interest Statement

The authors have declared no competing interest.

### Funding Statement

This study was funded by National Natural Science Foundation of China (No. 61827809) and the National Key Research and Development Program of China (No. 2020YFA0712200)

